# Clinical and immunological factors that distinguish COVID-19 from pandemic influenza A(H1N1)

**DOI:** 10.1101/2020.08.10.20170761

**Authors:** José Alberto Choreño-Parra, Luis Armando Jiménez-Álvarez, Alfredo Cruz-Lagunas, Tatiana Sofía Rodríguez-Reyna, Gustavo Ramírez-Martínez, Montserrat Sandoval-Vega, Diana Lizzeth Hernández-García, Eduardo M. Choreño-Parra, Yalbi I. Balderas-Martínez, Mariana Esther Martinez-Sanchez, Eduardo Márquez-García, Edda Shiutto, José Moreno-Rodríguez, José Omar Barreto-Rodríguez, Hazel Vázquez-Rojas, Gustavo Iván Centeno-Sáenz, Néstor Alvarado-Peña, Citlaltepetl Salinas-Lara, Carlos Sanchez-Garibay, Gabriela Hernández, Criselda Mendoza-Milla, Andrea Domínguez, Julio Granados, Lula Mena-Hernández, Luis Ángel Pérez-Buenfil, Guillermo Domínguez-Cheritt, Carlos Cabello-Gutiérrez, Cesar Luna, Jorge Salas-Hernández, Patricio Santillán-Doherty, Justino Regalado, Angélica Hernández-Martínez, Lorena Orozco, Ethel Awilda García-Latorre, Carmen M. Hernández-Cárdenas, Shabaana A. Khader, Albert Zlotnik, Joaquín Zúñiga

**Affiliations:** Escuela Nacional de Ciencias Biológicas, Instituto Politécnico Nacional, Mexico City, Mexico; Laboratory of Immunobiology and Genetics, Instituto Nacional de Enfermedades Respiratorias Ismael Cosío Villegas, Mexico City, Mexico; Department of Immunology and Rheumatology, Instituto Nacional de Ciencias Médicas y Nutrición Salvador Zubirán, Mexico City, Mexico; Facultad de Estudios Superiores Iztacala, Universidad Nacional Autónoma de México, Mexico City, Mexico; Intensive Care Unit, Instituto Nacional de Enfermedades Respiratorias Ismael Cosío Villegas, Mexico City, Mexico; Posgrado en Ciencias Biológicas, Universidad Nacional Autónoma de México, Mexico City, Mexico; Department of Immunology, Instituto de Investigaciones Biomédicas, Universidad Nacional Autónoma de México, Mexico City, Mexico; Direccion de Enseñanza e Investigación, Hospital Juárez de Mexico, Mexico City, Mexico; Subdirección de Medicina y, Instituto Nacional de Enfermedades Respiratorias Ismael Cosío Villegas, Mexico City, Mexico; Coordinación de Infectología y Microbiología, Instituto Nacional de Enfermedades Respiratorias Ismael Cosío Villegas, Mexico City, Mexico; Departamento de Neuropatología, Instituto Nacional de Neurología y Neurocirugía “Manuel Velasco Suarez”, Mexico City, Mexico; Tecnologico de Monterrey, Escuela de Medicina y Ciencias de la Salud, Mexico City, Mexico; Department of Transplantation, Instituto Nacional de Ciencias Médicas y Nutrición Salvador Zubirán, Mexico City, Mexico; Department of Dermatology, Instituto Nacional de Ciencias Médicas y Nutrición Salvador Zubirán, Mexico City, Mexico; Department of Education, Instituto Nacional de Ciencias Médicas y Nutrición Salvador Zubirán, Mexico City, Mexico; Department of Critical Care Unit, Instituto Nacional de Ciencias Médicas y Nutrición Salvador Zubirán, Mexico City, Mexico; Department of Virology, Instituto Nacional de Enfermedades Respiratorias Ismael Cosío Villegas, Mexico City, Mexico; Department of Pathology, Instituto Nacional de Enfermedades Respiratorias Ismael Cosío Villegas, Mexico City, Mexico; Department of General Direction, Instituto Nacional de Enfermedades Respiratorias Ismael Cosío Villegas, Mexico City, Mexico; Department of Medical Direction, Instituto Nacional de Enfermedades Respiratorias Ismael Cosío Villegas, Mexico City, Mexico; Instituto Nacional de Medicina Genómica, Mexico City, Mexico; Department of Molecular Microbiology, Washington University School of Medicine in St Louis, MO, United States; Department of Physiology & Biophysics School of Medicine, Institute for Immunology, University of California, Irvine, United-States

**Author notes:** **Correspondence: Joaquín Zúñiga PhD.** Laboratory of Immunobiology and Genetics, Instituto Nacional de Enfermedades Respiratorias Ismael Cosío Villegas, Mexico City, Mexico.

## Abstract

The severe acute respiratory syndrome coronavirus 2 (SARS-CoV-2), the causative agent of coronavirus disease 2019 (COVID-19), is a global health threat with the potential to cause severe disease manifestations in the lungs. Although clinical descriptions of COVID-19 are currently available, the factors distinguishing SARS-CoV-2 from other respiratory viruses are unknown. Here, we compared the clinical, histopathological, and immunological characteristics of patients with COVID-19 and pandemic influenza A(H1N1). We observed a higher frequency of respiratory symptoms, increased tissue injury markers, a histological pattern of alveolar pneumonia, and higher levels of IL-1RA, TNF-α, CCL3, G-CSF, APRIL, sTNF-R1, sTNF-R2, sCD30, and sCD163 in influenza patients. Conversely, dry cough, gastrointestinal symptoms, interstitial lung pathology, increased Th1 (IL-12, IFN-γ) and Th2 (IL-4, IL-5, IL-10, IL-13) cytokine levels, along with IL-1β, IL-6, CCL11, VEGF, TWEAK, TSLP, MMP-1, and MMP-3, were observed in COVID-19 cases. We demonstrated the diagnostic potential of some clinical and immune factors to differentiate COVID-19 from pandemic influenza A(H1N1). Our data suggest that SARS-CoV-2 induces a dysbalanced polyfunctional inflammatory response that is different from the immune response against influenza. These findings might be relevant for the upcoming 2020-2021 influenza season, which is projected to be historically unique due to its convergence with COVID-19.

## Introduction

The novel SARS-CoV-2 has submerged the world into a public health crisis of unprecedented features. With more than 18.6 million infected people and 702,000 deaths, SARS-CoV-2 continues spreading worldwide (1). Although other emerging pathogens have caused similar outbreaks in the past, the pandemic influenza A(H1N1) pdm09 virus is the immediate antecedent reference for the global spread of a new zoonotic respiratory pathogen. This virus emerged in Mexico in 2009, causing approximately 151,700-575,400 deaths worldwide during the first year after its appearance (2-4). Ever since, the influenza A(H1N1) pdm09 virus has continued circulating globally, acquiring a seasonal transmission pattern (5). Notably, the emergence of SARS-CoV-2 in December of 2019 (6-8), occurred when several countries were at the peak of the flu season. This hampered differentiating COVID-19 and influenza during the early days of the current pandemic. With improved understanding of the clinical COVID-19 pathobiology (9-12), the overall identification of COVID-19 positive cases has improved.

Despite this, only a few comparisons of the characteristics of COVID-19 and influenza have been recently conducted (13-15). This is crucial as it is highly likely that both entities will converge during the next influenza season in the Northern hemisphere. During such a predicted scenario, the accurate identification of the causative pathogen may have important therapeutic implications, including the selection of adequate antiviral drugs. A better understanding of the host factors implicated in protective vs. pathogenic immunity against SARS-CoV-2 is also crucial to guide immunotherapeutic interventions for patients in critical conditions. Thus, comparing the clinical and immune profiles of these two infectious entities may allow dissipating the prevailing controversies about the immune mechanisms mediating lung injury and mortality after SARS-CoV-2 infection by identifying unique immune signatures in COVID-19 but not influenza patients.

Here, we evaluated clinical and immunological factors distinguishing critically ill COVID-19 and pandemic influenza A(H1N1) patients. We also compared histopathological changes and expression of immune markers in the lungs of patients with SARS-CoV-2 and influenza A(H1N1) pdm09 virus infection. Our results reveal crucial differences in the clinical characteristics of both diseases. Furthermore, our analyses clearly show that the human immune response elicited after SARS-CoV-2 is completely different from the immune responses against the influenza A(H1N1) pdm09 virus. Our study provides evidence in favor of using some of these distinctive traits to differentiate SARS-CoV-2 infection from influenza at the hospital setting reliably.

## Results

### Clinical characteristics of influenza and COVID-19 patients

We retrieved clinical data and collected serum/plasma samples from patients with laboratory-confirmed COVID-19 that attended two national reference institutes of health in Mexico City (see Methods). This cohort included subjects admitted to the hospital that did not require mechanical ventilation (MV; moderate COVID-19) and individuals that required MV and admission to the intensive care unit (ICU; severe COVID-19). Our comparative cohort consisted of individuals with influenza A(H1N1) pdm09 virus infection (hereinafter referred to as influenza), requiring intubation and ICU care during the 2019-2020 influenza season immediately preceding the outbreak of COVID-19 in Mexico.

The main demographic characteristics of enrolled patients were similar (Table 1), although the proportion of males tended to be higher in both groups of COVID-19 subjects, as reported before (9, 10, 12, 16-18). Obesity was more frequent in influenza patients, whereas other comorbidities (diabetes, systemic arterial hypertension (SAH), chronic obstructive pulmonary disease (COPD), and obstructive sleep apnea syndrome (OSA)) were equally distributed across groups. Fever was the most frequent symptom among all participants, followed by cough, fatigue, myalgia, arthralgia, and headache. Dyspnea was present in 10% of patients with moderate COVID-19 and ~80% of individuals with severe COVID-19 and influenza. Rhinorrhea, sore throat, thoracic pain, and sputum production were more common during influenza, whereas dry cough, diarrhea, and vomit were more frequent among COVID-19 patients, suggesting that some symptoms could differentiate these infectious entities. We performed a logistic regression analysis with the symptoms reported by influenza and COVID-19 patients at hospital admission. Fever and rhinorrhea were associated with influenza, whereas dry cough predicted COVID-19 (Supplemental Figure 1 and Supplemental Table 2). Sore throat and thoracic pain were marginally associated with influenza but did not reach statistical significance. Similarly, gastrointestinal symptoms exhibited higher, but not significant odds ratio (OR) values for SARS-CoV-2 infection (Supplemental Figure 1 and Supplemental Table 2). Overall, patients in the moderate COVID-19 group attended earlier after illness onset than individuals with severe influenza and COVID-19 (Table 1).

**Table 1.** Clinical characteristics of patients with COVID-19 and influenza.

|  |  |  |  |  |  |  |
| --- | --- | --- | --- | --- | --- | --- |
| **Characteristic** | **Influenza**<br>**A(H1N1) pdm09** |  | **Moderate**<br>**COVID-19** |  | **Severe**<br>**COVID-19** |  |
|  | **A**<br>**N = 23** | ***p*-value**<br>**A vs. B** | **B**<br>**N = 10** | ***p*-value**<br>**B vs. C** | **C**<br>**N = 24** | ***p*-value**<br>**A vs. C** |
| **Age (years), median (range)** | 49 (29 - 77) | 0.2385 | 34.5 (28 – 71) | 0.1706 | 52 (30 – 73) | >0.9999 |
| **Gender** |  |  |  |  |  |  |
| Males | 14 (60.86) | 0.7098 | 7 (70) | 0.5659 | 19 (79.16) | 0.1703 |
| Females | 9 (39.13) |  | 3 (30) |  | 5 (20.83) |  |
| **BMI** | 33.6 (29.6 - 42.4) | 0.0004 | 25.3 (22.5 – 29.3) | 0.1164 | 29.6 (25.3 – 33.4) | 0.0592 |
| **Relevant co-morbidities** |  |  |  |  |  |  |
| Smoking | 8 (34.78) | 0.0715 | 0 (0) | 0.0720 | 8 (33.33) | 0.9165 |
| Biomass exposure | 5 (21.73) | 0.2911 | 0 (0) | 0.2958 | 4 (16.66) | 0.7238 |
| Diabetes | 5 (21.73) | >0.9999 | 2 (20) | >0.9999 | 6 (25) | 0.7918 |
| SAH | 6 (26.08) | >0.9999 | 2 (20) | >0.9999 | 4 (16.66) | 0.4936 |
| OSA | 2 (8.69) | >0.9999 | 0 (0) | 1 | 0 (0) | 0.2340 |
| COPD | 2 (8.69) | >0.9999 | 1 (10) | 0.2941 | 0 (0) | 0.2340 |
| Cancer | 0 (0) | 0.0852 | 2 (20) | 0.0802 | 0 (0) | 1 |
| **Clinical findings at onset** |  |  |  |  |  |  |
| Fever | 21 (91.3) | 0.3605 | 8 (80) | 0.7541 | 18 (75) | 0.1371 |
| Myalgia | 17 (73.91) | 0.7077 | 8 (80) | 0.5809 | 17 (70.83) | 0.8135 |
| Arthralgia | 17 (73.91) | 0.4438 | 6 (60) | 0.7109 | 16 (66.66) | 0.5871 |
| Headache | 11 (47.82) | 0.9086 | 5 (50) | 0.8245 | 11 (45.83) | 0.8911 |
| Dyspnea | 18 (78.26) | 0.0004 | 1 (10) | 0.0002 | 19 (79.16) | 0.9395 |
| Nasal congestion | 3 (13.04) | 0.5363 | 0 (0) | >0.9999 | 2 (8.33) | 0.6662 |
| Rhinorrhea | 11 (47.82) | 0.0129 | 0 (0) | 0.1478 | 6 (25) | 0.1035 |
| Sore throat | 8 (34.78) | 0.0321 | 0 (0) | 0.1478 | 6 (25) | 0.4635 |
| Thoracic pain | 4 (17.39) | 0.2890 | 0 (0) | 1 | 0 (0) | 0.0496 |
| Cough | 19 (82.6) | 0.4155 | 7 (70) | 0.2226 | 21 (87.5) | 0.6378 |
| Sputum | 11 (47.82) | 0.0129 | 0 (0) | 0.2958 | 4 (16.66) | 0.0220 |
| Dry cough | 8 (34.78) | 0.0619 | 7 (70) | 0.9612 | 17 (70.83) | 0.0133 |
| Fatigue | 19 (82.6) | >0.9999 | 8 (80) | 0.9563 | 19 (79.16) | 0.7643 |
| Diarrhea | 2 (8.69) | 0.1493 | 3 (30) | 0.3943 | 4 (16.66) | 0.6662 |
| Nausea | 2 (8.69) | 0.1493 | 3 (30) | 0.1380 | 2 (8.33) | >0.9999 |
| Vomit | 0 (0) | 0.0220 | 3 (30) | 0.3284 | 3 (12.5) | 0.0797 |
| **Illness onset - hospital admission (days)** | 7 (4 - 8.5) | 0.0583 | 3 (0 - 5.7) | 0.0158 | 6 (5 - 13.2) | >0.9999 |
| **Vital signs at admission** |  |  |  |  |  |  |
| Body temperature (°C) | 37 (36.8 - 37) | 0.5195 | 36.5 (36.3 - 37.2) | 0.02 | 37 (37 - 37.7) | 0.2878 |
| Respiratory rate (bpm) | 26 (22 - 30) | 0.0018 | 20 (16.7 - 21.7) | 0.0518 | 24 (22 - 26) | 0.48 |
| Heart rate (bpm) | 93 (80 - 103) | >0.9999 | 90 (75.7 - 99.7) | 0.7698 | 84 (72 - 90) | 0.1295 |
| MAP (mmHg) | 82 (73.5 - 94.8) | >0.9999 | 87 (80.7 - 88.7) | 0.1820 | 75 (70.2 - 84.5) | 0.3202 |

Data are displayed as n (%) or median (IQR). N is the total number of patients with available data. BMI, body mass index; bpm, breaths/beats per minute; COPD, chronic obstructive pulmonary disease; IQR, interquartile range; ICU, intensive care unit; MAP, mean arterial pressure; OSA, obstructive sleep apnea syndrome; SAH, systemic arterial hypertension; SD, standard deviation. Differences in continuous variables were estimated using the Kruskal Wallis with post hoc Dunn's test. Differences in categorical variables were calculated using the Fisher's exact or the Chi-square test as appropriate.

### Laboratory parameters that differentiate influenza from COVID-19

White blood cells (WBC), neutrophil counts, neutrophil to lymphocyte ratio (NLR), glucose, total bilirubin, and aspartate aminotransferase (AST) levels were similar in both influenza and severe COVID-19 groups, but lower in the moderate COVID-19 group (Table 2). Low lymphocyte counts were observed among all participants, indicating that lymphopenia is not a unique feature of severe COVID-19. Renal function parameters did not differ between groups. However, levels of some tissue injury markers, such as alkaline phosphatase (ALP), alanine aminotransferase (ALT), lactate dehydrogenase (LDH), creatine phosphokinase (CPK), and procalcitonin, were higher in influenza as compared to COVID-19 patients. We also observed that the Sequential Organ Failure Assessment (SOFA) and the Acute Physiology and Chronic Health Evaluation II (APACHE II) scores were higher in influenza patients. Despite this, the mortality of our cohort of critically ill influenza patients was significantly lower (21%) than the mortality of severely ill COVID-19 patients (62%), even when both groups presented similar rates of complications, and received equal supportive medical interventions (Table 3). No fatality cases were observed in the group of moderated COVID-19.

**Table 2.** Laboratory parameters of participants at admission.

| Parameter | Influenza<br>A(H1N1) pdm09<br>A<br>N = 23 | *p*-value<br>A vs. B | Moderate<br>COVID-19<br>B<br>N = 10 | *p*-value<br>B vs. C | Severe<br>COVID-19<br>C<br>N = 24 | *p*-value<br>A vs. C |
| --- | --- | --- | --- | --- | --- | --- |
| **Glucose (mg/dL)** | 132.1 (111 – 207.4) | 0.1241 | 96.5 (85.2 – 112) | 0.3025 | 124.3 (99 – 163.7) | >0.9999 |
| **Blood count** |  |  |  |  |  |  |
| White blood cells (10<sup>9</sup>/L) | 7.3 (5.8 – 11.9) | 0.0069 | 4.0 (3.5 – 5.7) | 0.0007 | 9.5 (6.4 – 13.1) | >0.9999 |
| Neutrophils (10<sup>9</sup>/L) | 5.7 (4.6 – 9.9) | 0.0057 | 2.9 (1.8 – 3.9) | 0.0184 | 7.4 (4.1 – 10.1) | >0.9999 |
| Lymphocytes (10<sup>9</sup>/L) | 0.7 (0.4 – 0.9) | 0.5050 | 0.9 (0.6 – 1.2) | >0.9999 | 0.9 (0.6 – 1.2) | 0.3522 |
| NLR | 8.4 (5.1 – 17.1) | 0.0120 | 3.1 (1.6 – 5.7) | 0.0702 | 8.7 (3.7 – 13.4) | >0.9999 |
| Hgb (g/dL) | 14.2 (13 – 17.3) | 0.6994 | 15.4 (14.2 – 16.4) | 0.0755 | 13.7 (13.2 – 15.1) | 0.5663 |
| Platelets (10<sup>9</sup>/L) | 173 (141 – 205) | >0.9999 | 192 (137 – 224) | 0.7794 | 208 (165 – 258) | 0.1547 |
| **Renal function** |  |  |  |  |  |  |
| Cr (mg/dL) | 1.1 (0.9 – 2.2) | 0.5773 | 1.0 (0.8 – 1.1) | >0.9999 | 0.9 (0.8 – 1.5) | 0.4344 |
| BUN (mg/dL) | 22.2 (16.2 – 34.3) | 0.1106 | 14.5 (10.5 – 18.8) | 0.6716 | 18 (11.9 – 26.8) | 0.7558 |
| Na (mmol/L) | 135.2 (132.5 – 139.3) | >0.9999 | 137 (136 – 139) | 0.4082 | 139.8 (136.2 – 141.7) | 0.0161 |
| K (mmol/L) | 4.2 (3.8 – 4.6) | 0.7951 | 4.0 (3.7 – 4.2) | 0.6386 | 4.2 (4 – 4.5) | >0.9999 |
| **Liver function** |  |  |  |  |  |  |
| Total bilirubin (mg/dL) | 0.6 (0.4 – 0.8) | 0.0344 | 0.3 (0.3 – 0.4) | 0.0183 | 0.5 (0.4 – 0.8) | >0.9999 |
| AST (U/L) | 60.9 (39.6 – 84.1) | 0.0026 | 22.6 (15 – 38.5) | 0.0170 | 43.5 (29 – 90.7) | >0.9999 |
| ALT (U/L) | 29.2 (23.3 – 47.5) | 0.4934 | 21.7 (17.2 – 32.1) | 0.0435 | 40.2 (28.8 – 56.8) | 0.5891 |
| ALP (U/L) | 122.7 (86.1 – 169.7) | 0.0016 | 72.5 (58.2 – 80.5) | 0.6821 | 78 (63.4 – 88.2) | 0.0104 |
| **Other biomarkers** |  |  |  |  |  |  |
| LDH (U/L) | 643.8 (452.2 – 804.7) | <0.0001 | 186 (165.8 – 251.5) | 0.0070 | 414.5 (318.4 – 494.8) | 0.0269 |
| CPK (U/L) | 274.4 (158.2 – 771.2) | 0.0277 | 73 (49.7 – 161.3) | 0.1225 | 160.3 (74.6 – 1419) | >0.9999 |
| Procalcitonin (ng/mL) | 0.6 (0.2 – 3.6) | <0.0001 | 0.05 (0.05 – 0.08) | 0.1652 | 0.1 (0.09 – 0.17) | 0.0008 |
| **Gasometric parameters** |  |  |  |  |  |  |
| pH | 7.37 (7.32 – 7.45) | 0.2325 | 7.43 (7.41 – 7.46) | 0.3277 | 7.41 (7.33 – 7.45) | >0.9999 |
| PaO<sub>2</sub> mmHg | 51 (38 – 67) | 0.0768 | 65 (55 – 91) | 0.1260 | 51 (42 – 65) | >0.9999 |
| PCO<sub>2</sub> mmHg | 35 (30 – 48) | >0.9999 | 34 (29 – 37) | >0.9999 | 34 (27 – 47) | >0.9999 |
| Lactate (mmol/L) | 1.2 (0.8 – 1.5) | ND | ND | ND | 0.9 (0.8 – 1.1) | 0.1309 |
| HCO<sub>3</sub> (mEq/L) | 22.2 (17.9 – 27.4) | >0.9999 | 22.6 (21.5 – 25.8) | 0.4845 | 21.1 (19 – 22.8) | 0.9628 |
| **PaO<sub>2</sub>/FiO<sub>2</sub>** | 96 (62.8 – 160) | <0.0001 | 314 (262 – 433) | 0.001 | 127 (96 – 155) | 0.5999 |
| Mild (PaO<sub>2</sub>/FiO<sub>2</sub> 201 - 300) | 1 (4.34) | 0.0002 | 7 (70) | 0.0019 | 3 (12.5) | 0.6085 |
| Moderate (PaO<sub>2</sub>/FiO<sub>2</sub> 101-200) | 11 (47.82) | 0.3410 | 3 (30) | 0.0836 | 15 (62.5) | 0.3118 |
| Severe (PaO<sub>2</sub>/FiO<sub>2</sub> <100) | 11 (47.82) | 0.0129 | 0 (0) | 0.0815 | 6 (25) | 0.1035 |
| **Severity of illness scores** |  |  |  |  |  |  |
| SOFA | 8 (7 – 13) | <0.0001 | 1 (0 – 2) | 0.0031 | 6 (3 – 8) | 0.0398 |
| APACHE II | 11 (5 – 18) | 0.0405 | 4 (0 – 7.5) | 0.3546 | 7 (4 – 10) | 0.6420 |

Data are displayed as n (%) or median (IQR). N is the total number of patients with available data. ALP, alkaline phosphatase; APACHE-II, Acute Physiology and Chronic Health Evaluation II; AST, aspartate aminotransferase; ALT, alanine aminotransferase; BUN, blood ureic nitrogen; CPK, creatine phosphokinase; Cr, creatinine; FiO<sub>2</sub>, fraction of inspired oxygen; HCO<sub>3</sub>, bicarbonate; Hgb, hemoglobin; IQR, interquartile range; LDH, lactate dehydrogenase; ND, not determined; NLR, neutrophil/lymphocyte ration; PaO<sub>2</sub>, partial pressure of oxygen in arterial blood; PCO<sub>2</sub>, partial pressure of carbon dioxide in the blood; SD, standard deviation; SOFA, Sequential Organ Failure Assessment. Differences in continuous variables were estimated using the Kruskal Wallis with post hoc Dunn's test. Differences in categorical variables were calculated using the Fisher's exact or the Chi-square test as appropriate.

**Table 3.** Complications and treatment of study participants.

| Parameter | Influenza<br>A(H1N1) pdm09 | *p*-value<br>A vs. B | Moderate<br>COVID-19 | *p*-value<br>B vs. C | Severe<br>COVID-19 | *p*-value<br>A vs. C |
| --- | --- | --- | --- | --- | --- | --- |
|  | A<br>N = 23 |  | B<br>N = 10 |  | C<br>N = 24 |  |
| **Complications** |  |  |  |  |  |  |
| Acute myocardial infarction | 3 (13.04) | 0.5363 | 0 (0) | 1 | 0 (0) | 0.1092 |
| Deep vein thrombosis | 1 (4.34) | >0.9999 | 0 (0) | 1 | 0 (0) | 0.4894 |
| Acute kidney injury | 10 (43.47) | 0.0148 | 0 (0) | 0.0815 | 6 (25) | 0.1814 |
| Secondary infection | 14 (60.86) | 0.0014 | 0 (0) | 0.0169 | 10 (41.66) | 0.1880 |
| **Medical treatment** |  |  |  |  |  |  |
| Oseltamivir | 23 (100) | 0.0012 | 6 (60) | 0.9283 | 14 (58.33) | 0.0005 |
| Antibiotic therapy | 23 (100) | 1 | 10 (100) | 1 | 24 (100) | 1 |
| No. of antibiotics per patient, median (range) | 3.5 (2 – 10) | 0.0077 | 2 (2 – 3) | 0.0252 | 4 (3 – 5) | >0.9999 |
| Chloroquine/hydroxychloroquine | 0 (0) | <0.0001 | 10 (100) | <0.0001 | 23 (95.83) | <0.0001 |
| Azithromycin | 0 (0) | <0.0001 | 10 (100) | 0.0049 | 13 (54.16) | <0.0001 |
| Corticosteroids | 4 (17.39) | 0.2890 | 0 (0) | 0.2908 | 5 (20.83) | 0.7643 |
| **Respiratory support** |  |  |  |  |  |  |
| Nasal cannula | 0 (0) | <0.0001 | 10 (100) | <0.0001 | 0 (0) | 1 |
| MV | 23 (100) | <0.0001 | 0 (0) | <0.0001 | 24 (100) | 1 |
| Prone position | 14 (60.86) | 0.0014 | 0 (0) | 0.0135 | 11 (45.83) | 0.3017 |
| ECMO | 2 (8.69) | 0.3360 | 0 (0) | 1 | 0 (0) | 0.2340 |
| **Renal replacement therapy** | 6 (26.08) | 0.1445 | 0 (0) | 0.2908 | 5 (20.83) | 0.4252 |
| **Mortality** | 5 (21.73) | 0.2911 | 0 (0) | 0.0135 | 15 (62.5) | 0.0077 |

Data are displayed as n (%) or median (IQR). N is the total number of patients with available data. ECMO, extracorporeal membrane oxygenation; IQR, interquartile range; MV, mechanical ventilation; SD, standard deviation. Differences in continuous variables were estimated using the Kruskal Wallis with post hoc Dunn's test. Differences in categorical variables were calculated using the Fisher's exact or the Chi-square test as appropriate.

### Serum cytokine profiles in influenza and COVID-19 patients

The severity of influenza and COVID-19 has been systematically attributed to an exacerbated production of pro-inflammatory cytokines (cytokine storm syndrome (CSS)) (19, 20). More recently, some researchers have also proposed that a depression of the functional capacity of the immune system, rather than an exuberant immune activation, is responsible for the clinical pathology of severe COVID-19 (21). Comparing the immune responses elicited by SARS-CoV-2 and influenza A(H1N1) pdm09 virus may be more helpful in identifying unique immune mechanisms associated with morbidity and mortality in COVID-19. Thus, we determined the circulating levels of several immune mediators in influenza and COVID-19 patients and correlated such levels with clinical findings and disease outcomes. Our results showed that critically ill COVID-19 patients had increased levels of IL-1β, IL-1RA, IL-6, IL-9, and CXCL10, and lower levels of IL-2 and IL17A as compared to healthy volunteer donors (Figure 1 and Supplemental Figure 2). These findings are coincident with the immune profiles that were reported in Chinese patients with COVID-19 (9, 22, 23). Levels of pro-inflammatory (IFN-γ, IL-1β, IL-6, IL-9, IL-12p70, CCL11) and anti-inflammatory (IL-4, IL-5, IL-10, IL-13) cytokines, as well as VEGF, were higher in severely ill COVID-19 patients as compared to influenza subjects. In contrast, levels of IL-1RA, IL-2, TNF-α, CCL3, and G-CSF were more increased among influenza patients as compared to severe COVID-19 patients (Figure 1 and Supplemental Figure 2).

**Figure 1.**
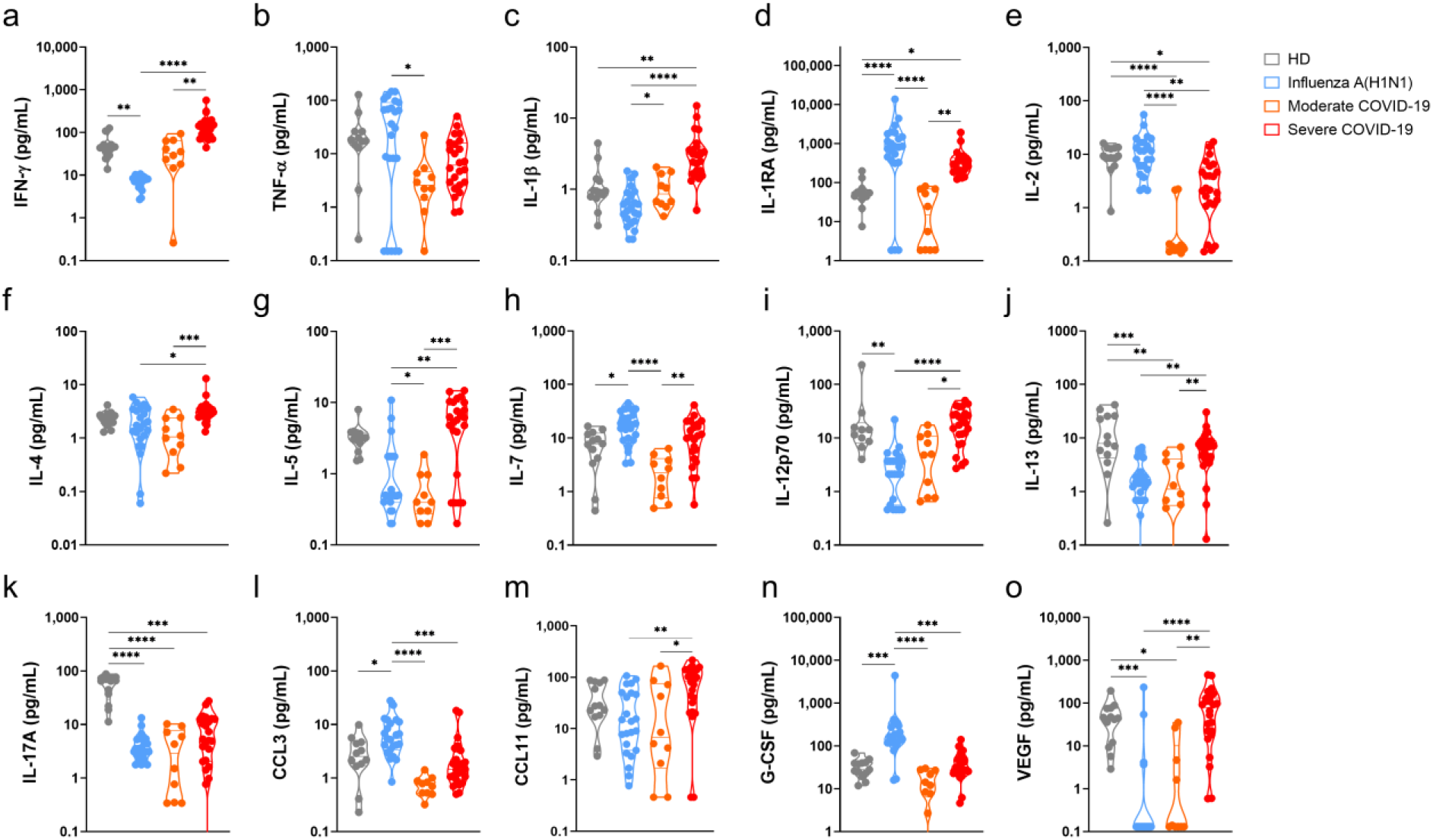
Serum cytokine levels in pandemic influenza A(H1N1) and COVID-19 patients. Serum levels of cytokines, chemokines, and growth factors in healthy volunteer donors (HD, n=13), patients with COVID-19 (n=10 moderate, 24 severe), and influenza (n=23), were assessed by Luminex assay. Violin plots display medians and interquartile ranges (IQR). Differences between groups we estimated using the Kruskal-Wallis test with post hoc Dunn’s test. Significant differences are denoted by bars and asterisks: *p≤0.05, **p≤0.01, ***p≤0.001, ****p≤0.0001. (a) IFN-γ, interferon-gamma; (b) TNF-α, tumor necrosis factor-alpha; (c) IL-1β, interleukin 1beta; (d) IL-1RA, interleukin 1 receptor antagonist; (e) IL-2, interleukin 2; (f) IL-4, interleukin 4; (g) IL-5, interleukin 5; (h) IL-7, interleukin 7; (i) IL-12p70, interleukin 12 p70 subunit; (j) IL-13, interleukin 13; (k) IL-17A, interleukin 17A; (l) CCL3, C-C motif chemokine ligand 3; (m) CCL11, C-C motif chemokine ligand 11; (n) G-CSF, granulocyte colony-stimulating factor; (o) VEGF, vascular endothelial growth factor.

These serum cytokine profiles indicate that, besides a higher production of pro-inflammatory and Th1 cytokines, SARS-CoV-2, but not influenza A(H1N1) pdm09 infection, parallelly induces Th2 responses. This may suggest that a lack of sufficient regulation and balancing of the type of immune response triggered after SARS-CoV-2 infection might contribute to the immune dysfunction reported during COVID-19. Also, the proinflammatory and profibrotic immune profile observed in COVID-19 patients may contribute to the extensive tissue damage and poor outcomes reported during SARS-CoV-2 infection. (21, 24) Other cytokines similarly increased in patients with severe influenza and COVID-19 included IL-7, IL-15, IL8, and CXCL10 (Supplemental Figure 2).

### Histological characteristics and expression of immune markers in the lungs of influenza and COVID-19 patients

Currently, no parallel comparative descriptions of the histological characteristics of the lung of patients with COVID-19 and influenza have been conducted. Here, we were able to obtain lung autopsy specimens from individuals that succumbed to either of these diseases and analyze their pathological features. Our histological analysis revealed that influenza induces alveolar edema and intra-alveolar inflammatory infiltrates in the lungs, which do not compromise the integrity of alveolar walls nor the micro-architecture of the organ (Figure 2a, left panel). These findings are compatible with a typical pattern of alveolar pneumonia. The inflammatory infiltrates observed in the lungs of influenza patients were composed of macrophages, polymorphonuclear cells, and scarce lymphocytes scattered between areas of intra-alveolar edema, hemorrhage, and fibrin mucoid exudates. Furthermore, although conserved, the alveolar walls showed capillaries with vasodilation and congestion (Figure 2b, left panel).

**Figure 2.**
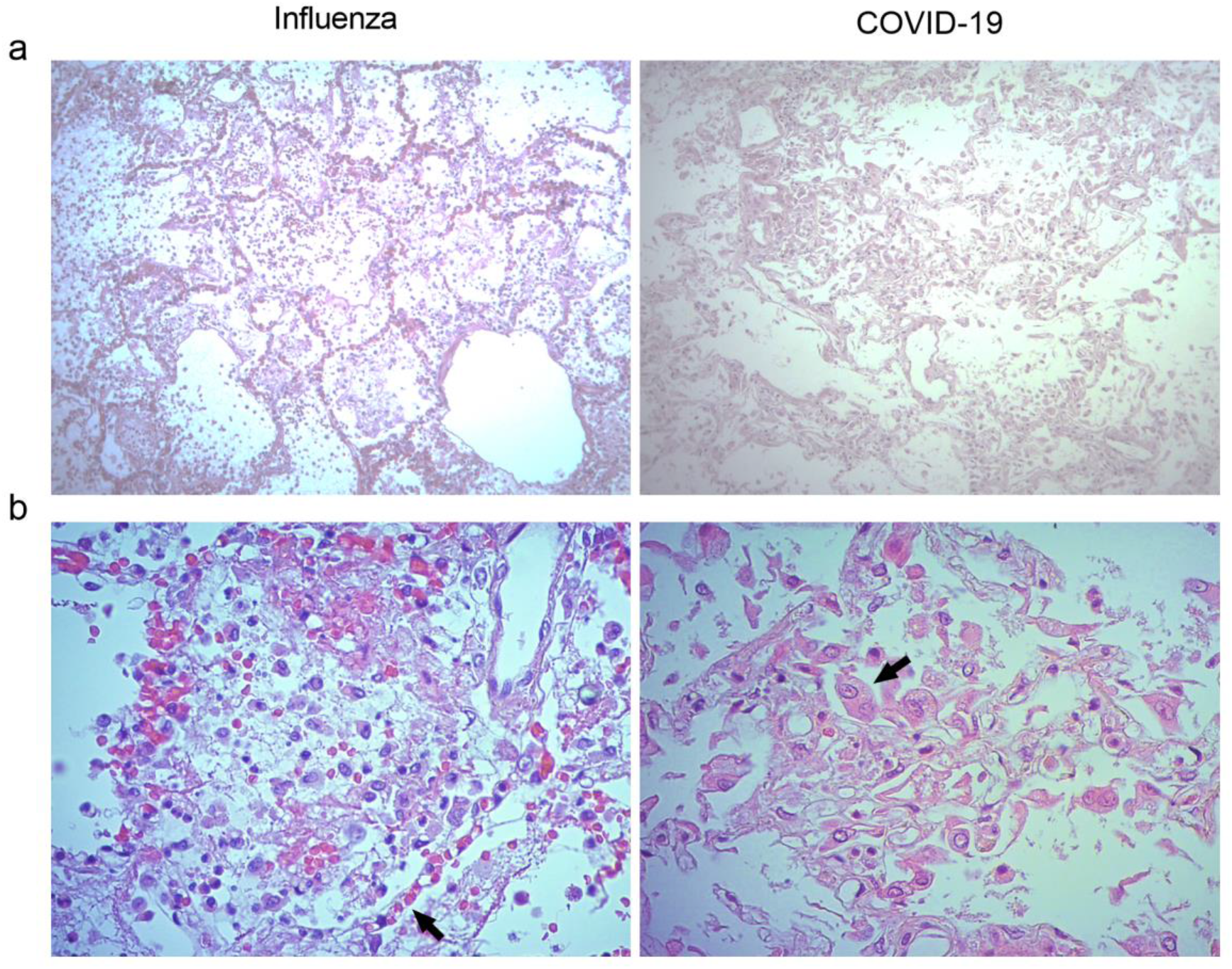
Histological characteristics of the lungs of patients with pandemic influenza A(H1N1) and COVID-19. Lung tissue autopsy specimens were obtained from patients that died of influenza and COVID-19. (a) The histological changes induced in the lungs during influenza were mainly characterized by intra-alveolar inflammatory infiltrates that did not compromise the integrity of alveolar walls (left panel). Meanwhile, the morphological changes of COVID-19 consisted of extensive inflammation, thickening of the alveolar walls, and partial loss of the histological architecture (right panel). H&E staining, x100. (b) The inflammatory infiltrates observed in the lungs of influenza patients consisted of macrophages, polymorphonuclear cells, and scarce lymphocytes scattered between areas of edema, hemorrhage, and fibrin deposits. Also, congestive, and vasodilated capillaries (arrow) were observed in the alveolar walls of influenza patients (left panel). Conversely, the inflammatory infiltrates found in the lung of COVID-19 patients were dominated by macrophages. Furthermore, the detachment of alveolar epithelial cells, which showed atypical characteristics such as large nucleoli (arrow), was also notable in COVID-19 patients (right panel). H&E staining, x400.

Meanwhile, SARS-CoV-2 induced distinctive morphological changes in the infected lung, characterized by an intense inflammatory infiltrate affecting extensive areas of the parenchyma, as well as a notable thickness of alveolar walls, hemorrhages, and partial loss of the histological architecture of the lung. These changes are compatible with interstitial pneumonia (Figure 2a, right panel). The inflammatory infiltrates observed in the lungs of COVID-19 patients were mainly composed of macrophages. A notable characteristic of the lungs infected with SARS-CoV-2 was the absence of lymphocytes and the detachment of pneumocytes, which showed hyperplasia, cellular changes, and prominent nucleoli (Figure 2b, right panel).

We also evaluated the expression of some immune markers in the lungs of influenza and COVID-19 patients by immunohistochemistry (IHQ). We found that IFN-γ, IL-1β, and IL-17A were expressed in the lungs of both groups of patients, mainly inside macrophages and pneumocytes (Figure 3). However, the intensity of expression of IFN-γ and IL-17A was higher in patients infected with SARS-CoV-2 as compared to influenza subjects. Strikingly, IL-4, which is a Th2 cytokine, was absent in the lungs of influenza patients but expressed in COVID-19 subjects (Figure 3). These findings are in line with the combined Th1/Th2 immune profile detected only in the serum of our cohort of patients infected with SARS-CoV-2 but not in influenza subjects.

**Figure 3.**
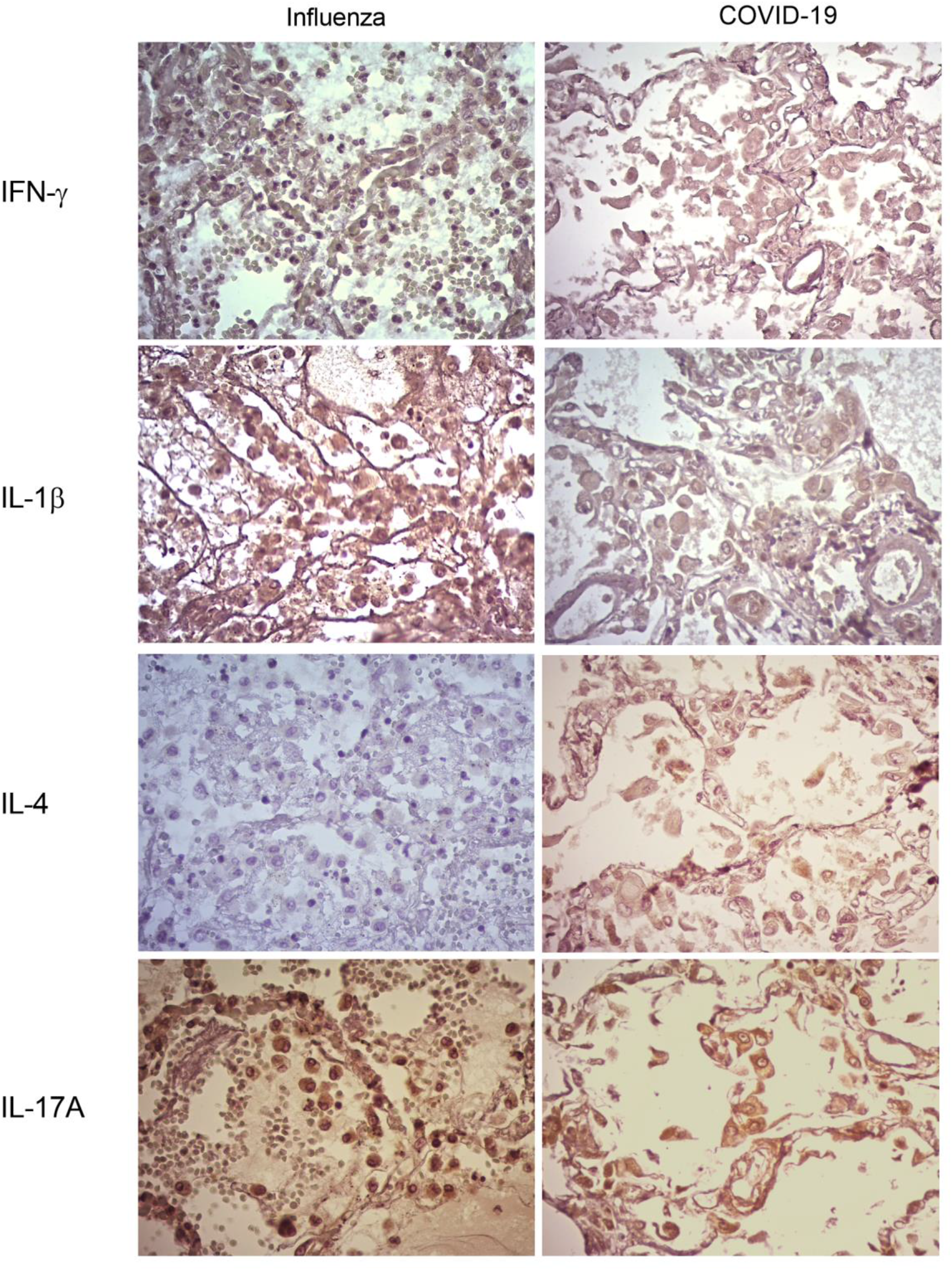
Expression of immune markers in the lungs of influenza and COVID-19 patients. Expression of different immune markers in lung autopsy specimens from influenza and COVID-19 patients was assessed using specific antibodies by immunohistochemistry (IHQ), x400.

### Clinical features, laboratory parameters, and immune markers that distinguish influenza and COVID-19

To determine which clinical and immunological characteristics contributed more to the differences between influenza and COVID-19, we performed a principal component analysis (PCA). The analysis showed that influenza patients cluster apart from the combined cohort of COVID-19 subjects in the PC2 (Figure 4a). Of note, clinical characteristics contributed to 31.2% of the total variance explained by the two first PCs (12.51% to PC1 and 50.03% to PC2). Meanwhile, serum cytokine levels contributed to 68.7% of the total variance explained by the two first PCs (87.48% to PC1 and 49.96% to PC2). These data indicate that immunological characteristics may be more useful than clinical variables to discriminate between influenza and COVID-19. Thus, we performed additional PCAs using only clinical or immunological characteristics. We observed that patients with severe influenza were not separated from severely ill COVID-19 patients by their clinical features, but they clustered apart from moderate COVID-19 subjects (Supplemental Figure 3a). Age, neutrophils, ALP, CPK, bilirubin, LDH, PaO2/FiO2, and SOFA were the clinical variables that contribute more to the first two PCs of this analysis. Conversely, influenza patients clustered apart from the entire COVID-19 cohort in a PCA using only serum cytokine levels (Supplemental Figure 3b). IFN-γ, IL-1RA, IL-5, IL-9, IL-10, and G-CSF levels contribute to the first two PCs of this PCA.

**Figure 4.**
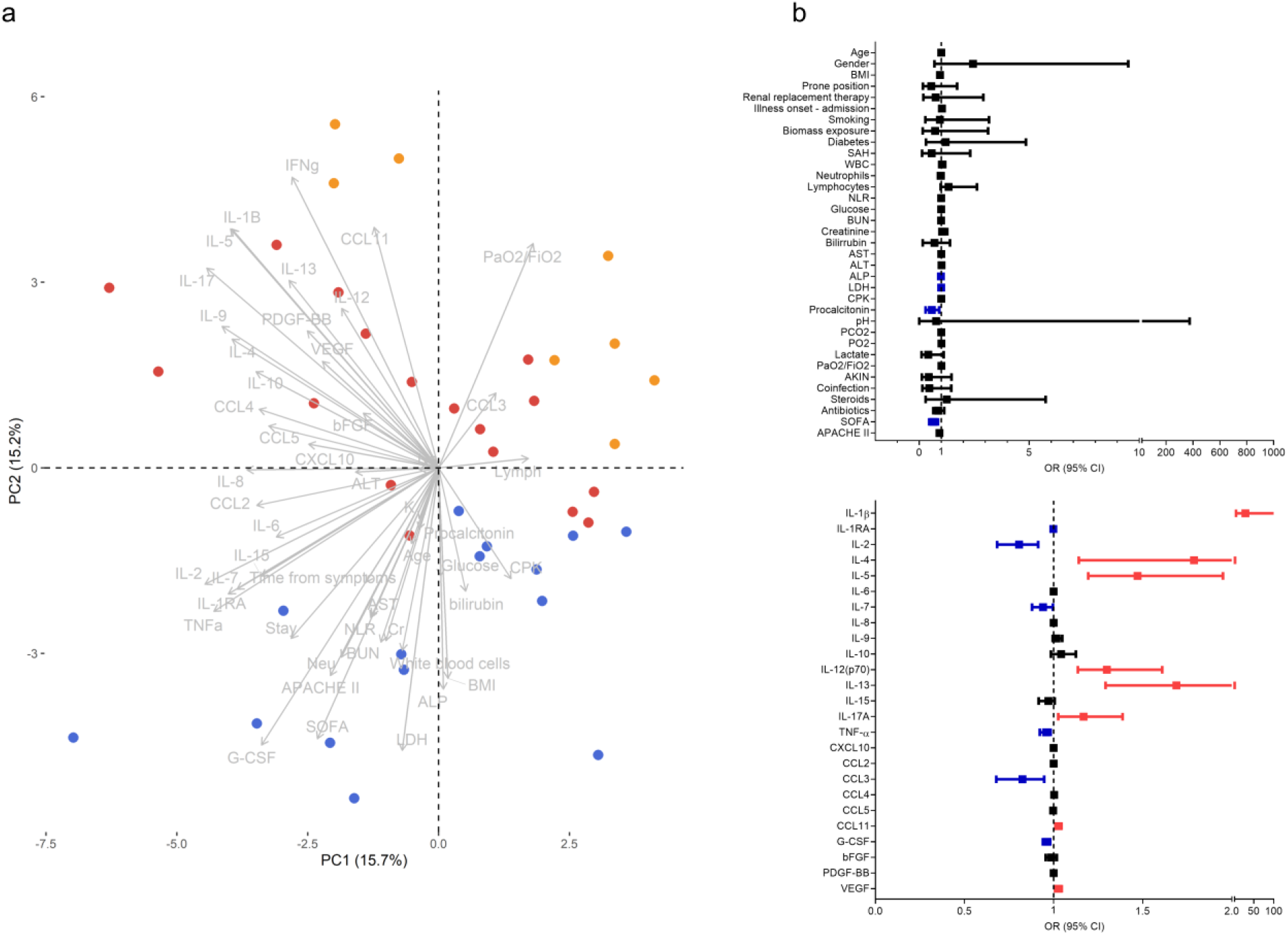
Clinical and immunological factors that distinguish pandemic influenza A(H1N1) and severe COVID-19. (a) Principal component analysis (PCA) of the clinical and immunological characteristics of study participants. Each dot represents a single individual, and each color represents a group of participants: blue for influenza A(H1N1), orange for moderate COVID-19, and red for severe COVID-19. (b) Bivariate logistic regression analysis of the clinical and immunological characteristics associated with the causative pathogen in the two cohorts of patients with severe influenza and COVID-19. The forest plots show the odds ratio (OR) and 95% CI interval values that were non-significant (black) and significant for severe COVID-19 (red). OR values of factors inversely associated with severe COVID-19 that instead predict influenza are shown in blue color. Absolute OR values are also presented in Supplementary Table 3.

Using logistic regression analyses, we further evaluated which clinical and immune factors differentiate our two cohorts of severely ill influenza and COVID-19 patients. IFN-γ was not included in this analysis, as it showed perfect discrimination of severe COVID-19 from influenza. We identified that LDH, ALP, procalcitonin, SOFA score, IL-1RA, IL-2, IL-7, TNF-α, CCL3, and G-CSF levels were significantly associated with severe influenza. In contrast, IL-1β, IL-4, IL-5, IL-12p70, IL-13, IL-17A, CCL11, and VEGF levels predicted severe COVID-19 (Figure 4b). Some of these factors, along with PaO2/FiO2 index, the incidence of acute kidney injury (AKIN), co-infections, APACHE-II score, IFN-γ, IL-15, and CCL5, also contributed to differentiate the entire cohort of patients with moderate and severe COVID-19 from influenza subjects (Supplemental Figure 4).

A linear discriminant analysis (LDA) showed that some of these selected parameters, along with AST and ALT, used together, accurately differentiate between severe influenza, moderate COVID-19, and severe COVID-19 groups (Figure 5a-b). Since it would be impractical to assess all these factors combined to differentiate both diseases, we analyze the results of the LDA using the Wilk’s Lambda test. This analysis showed that ALT, ALP, SOFA, IL-2, and TNF-α were crucial for the discriminative power of our LDA model (Figure 5c). Furthermore, receiver operating characteristics (ROC) curve analyses showed that IFN-γ, IL-1β, IL-12p70, G-CSF, and VEGF had the highest diagnostic performance to distinguish severe COVID-19 and influenza (Figure 6).

**Figure 5.**
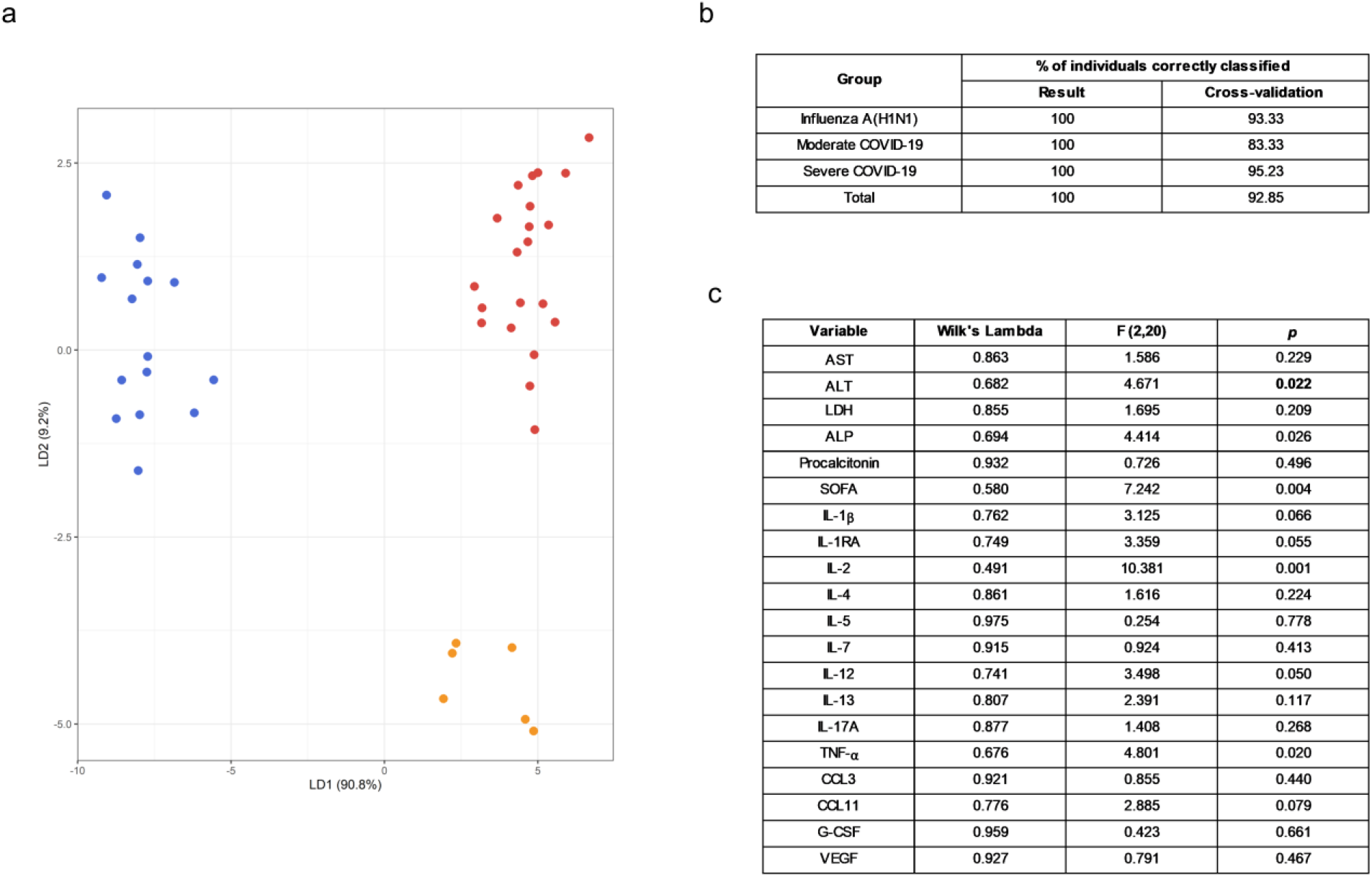
Selected clinical and immunological characteristics that better distinguish pandemic influenza A(H1N1) from COVID-19. (a) Linear discriminant analysis (LDA) plot of the first two discriminant functions showing the separation of the different groups of study participants according to a set of selected clinical and immunological characteristics used in combination. Each dot represents a single individual, and each color represents a group of participants: blue for influenza A(H1N1), orange for moderate COVID-19, and red for severe COVID-19. (b) Accuracy of the LDA results before and after a “leave-one-out” cross-validation. (c) The discriminant potential of each individual variable included in the LDA was estimated using the Wilk’s Lambda test. The table displays values of Wilk’s lambda, F, and *p* for each variable.

**Figure 6.**
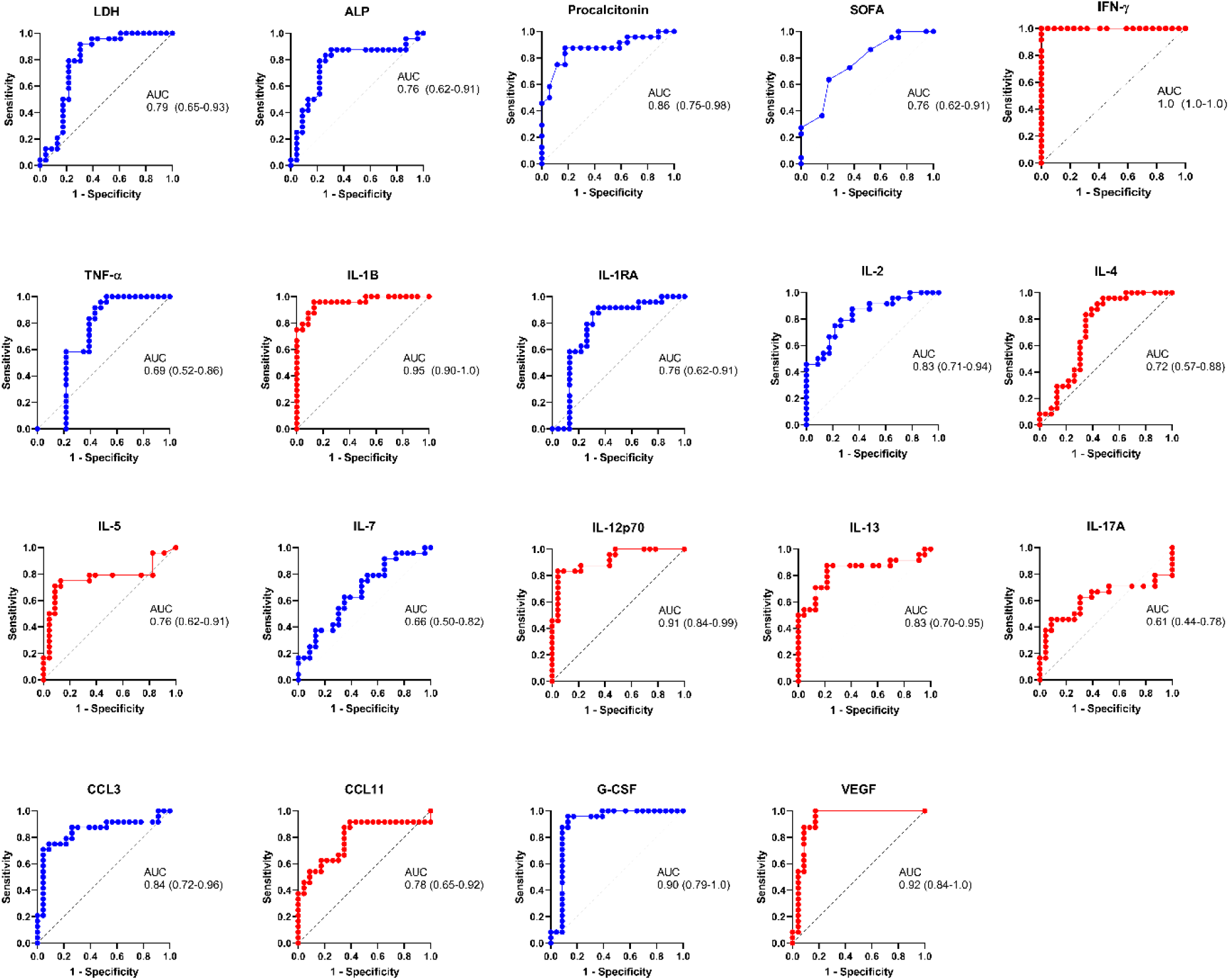
Diagnostic value of the clinical and immunological factors that distinguish between severe COVID-19 and influenza. Receiver operating characteristic (ROC) curves were constructed for the clinical and immunological characteristics that showed significant OR values in the bivariate logistic regression analysis. ROC curves of variables associated with influenza are shown in blue color, whereas variables associated with severe COVID-19 are displayed in red color. The graphs show area under the curve (AUC) and 95% CI interval values.

### Clinical and immunological factors associated with disease severity and outcome in influenza and COVID-19 patients

We also evaluated the prognostic value of clinical and immunological factors in influenza and COVID-19. Among COVID-19 patients, the duration of symptoms before admission, WBC, neutrophil counts, LDH, and SOFA score predicted severe disease defined as the need for intubation (Figure 7a). IL-4, IL-7, IL-8, IL-12p70, IL-15, and VEGF were also associated with increased risk of intubation in COVID-19 subjects. IL-6 showed increased but not significant OR values for severity in the combined COVID-19 cohort, contrasting with previous studies that indicate that IL-6 is significantly associated with severe COVID-19 (25, 26). Using a similar approach, we observed that WBC, and SOFA score conferred a higher risk of death after SARS-CoV-2 infection in the entire cohort of COVID-19 patients (Figure 7b).

**Figure 7.**
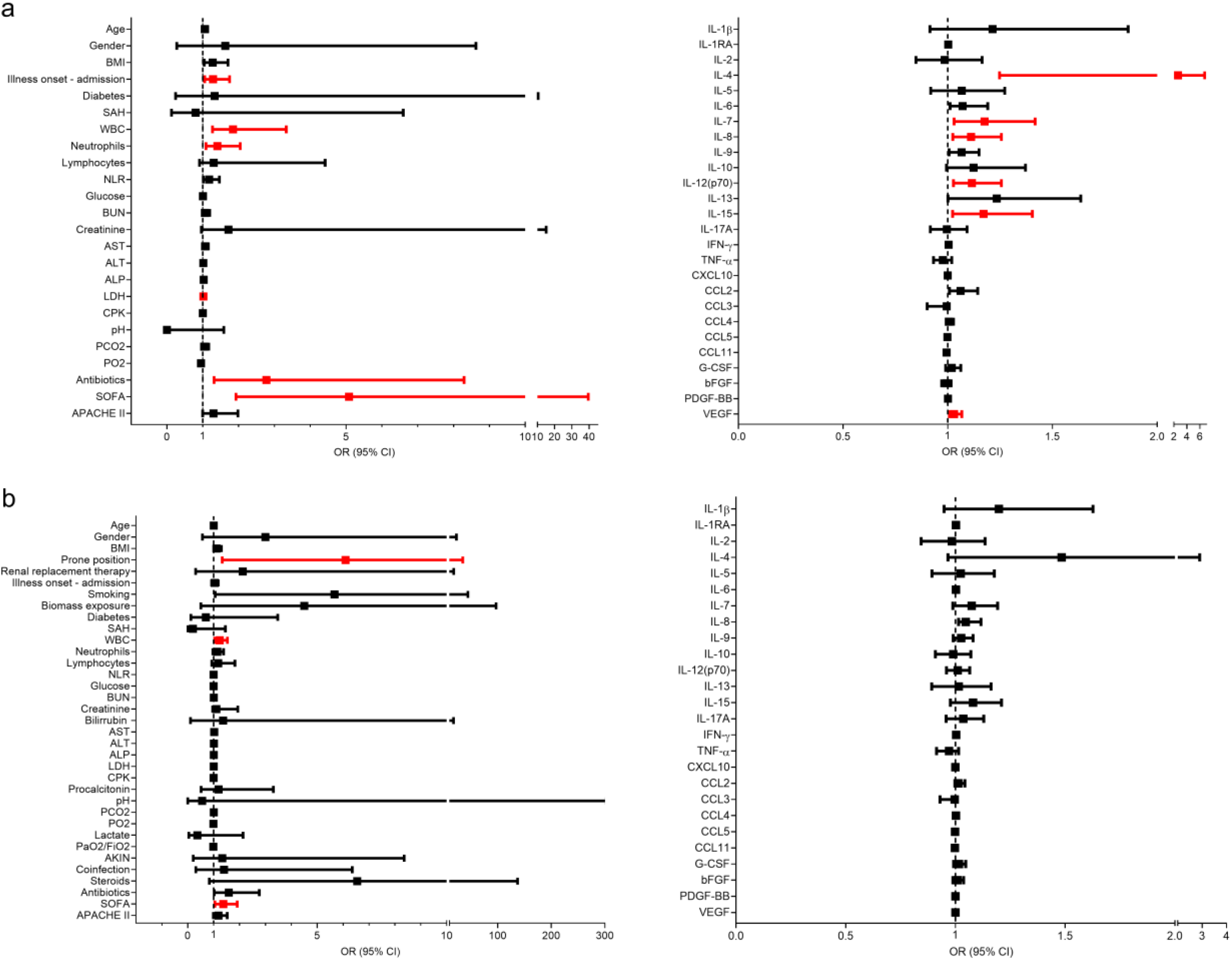
Clinical and immunological factors associated with disease outcomes in patients with COVID-19. (a) Bivariate logistic regression analysis of the clinical and immunological characteristics associated with intubation in patients COVID-19. (b) Clinical and immunological factors associated with mortality in patients with COVID-19. The forest plots show the odds ratio (OR) and 95% CI interval values. OR values that did not include the null value in the 95% CI were considered significant for intubation/mortality and are shown in red color. Absolute OR values are also presented in Supplemental Table 5.

Likewise, the need for renal replacement therapy (OR 32, 3 – 849.9 95% CI, *p* = 0.0029), and the use of steroids (OR 25.5, 2.1 – 698.4 95% CI, *p* = 0.0091), were associated with mortality risk after influenza (Supplemental Figure 5), as reported before (27, 28). However, none of the evaluated cytokines were associated with mortality in COVID-19 and influenza patients (Figure 7b and Supplemental Figure 5). At the time of patient recruitment, there was no consensus regarding the use of steroids for COVID-19, and the RECOVERY trial had not been published (29). Hence, only some of our COVID-19 patients were treated with steroids.

### Plasma levels of an extended set of immune markers that also distinguish influenza from COVID-19

Finally, we analyzed another set of immune mediators in the blood of 25 moderate and 24 severe COVID-19 patients, as well as in 22 influenza subjects, from which we were able to obtain plasma samples (Figure 8 and Supplemental Figure 6). Plasma levels of these factors showed only a few correlations with clinical characteristics and serum cytokine levels (Supplemental Figure 7). The overall profile of these correlations was different in influenza and COVID-19 patients, suggesting distinct immune mechanisms underlying clinical manifestations of both diseases.

**Figure 8.**
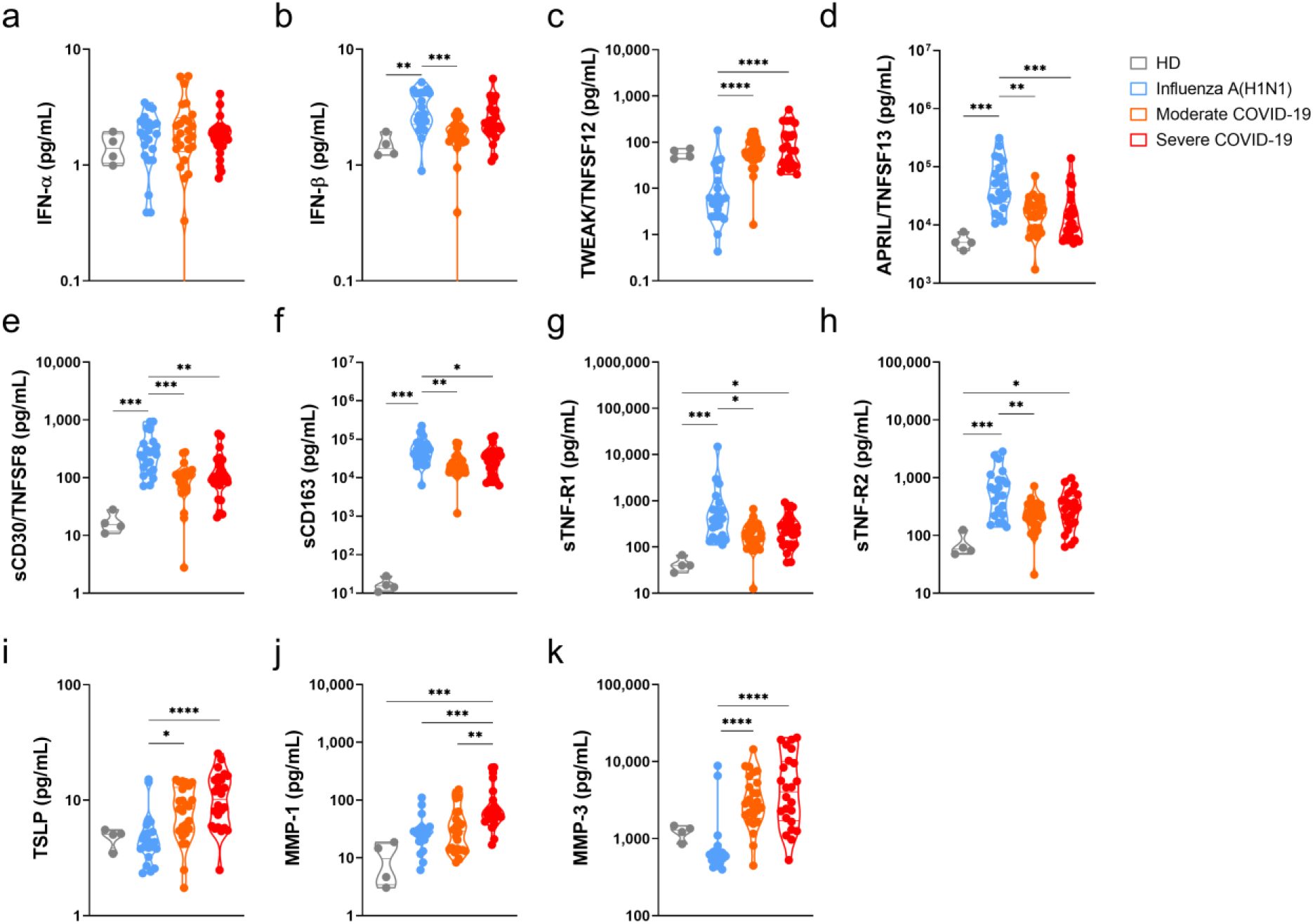
Immune mediators in the plasma of patients with pandemic influenza A(H1N1) and COVID-19. Levels of different soluble immune mediators in plasma samples from patients with COVID-19 (n=25 moderate, 24 severe) and influenza (n=23), as well as in samples from healthy volunteer donors (HD, n=4) were assessed by Luminex assay. Violin plots display medians and interquartile ranges (IQR). Differences between groups we estimated using the Kruskal-Wallis test with post hoc Dunn’s test. Significant differences are denoted by bars and asterisks: *p≤0.05, **p≤0.01, ***p≤0.001, ****p≤0.0001. (a) IFN-α, interferon-alpha; (b) IFN-β, interferon-beta; (c) TWEAK/TNFSF12, tumor necrosis factor-like weak inducer of apoptosis/tumor necrosis factor ligand superfamily member 12; (d) APRIL/TNFSF13, A proliferation-inducing ligand/tumor necrosis factor ligand superfamily member 13; (e) sCD30/TNFRSF8, soluble CD30/ tumor necrosis factor ligand superfamily member 8; (f) sCD163, soluble CD163; (g) sTNF-R1, soluble tumor necrosis factor receptor 1; (h) sTNF-R2, soluble tumor necrosis factor receptor 2; (i) TSLP, thymic stromal lymphopoietin; (j) MMP-1, metalloprotease 1; (k) MMP-3, metalloprotease 3.

Although levels of plasma type I interferons were below the levels of reliable detection, IFN-α, and IFN-β were increased among all participant groups as compared to healthy controls (Figure 8). Furthermore, a slight increase in the levels of IFN-β was noticed in influenza patients as compared to COVID-19 patients. Remarkably, although elevated, the levels of APRIL/TNFSF13, sCD30, sCD163, sTNF-R1, and sTNF-R2 were lower in COVID-19 than in influenza patients. APRIL/TNFSF13 is crucial for plasma cell survival (30). Thus, plasma cell responses could be downregulated in COVID—19 as compared to influenza. Soluble CD30 has been proposed as a marker of T cell activation during solid organ transplant rejection (31), whereas sCD163 is a readout of macrophage activation (32). Hence, our data may indicate a depletion of activated lymphocytes and macrophages from the circulation during SARS-CoV-2 infection, despite the high levels of inflammatory mediators found in COVID-19 patients. Soluble TNF-R1 and sTNF-R2 act as decoy receptors for TNF-α (33); as such, patients with COVID-19 might be less capable of balancing pathogenic TNF-α activities than individuals with influenza.

TWEAK, TSLP, MMP-1, and MMP-3 were elevated in COVID-19 cases. TWEAK is a stimulator of IL-6, IL-8, CXCL10, and MMP-1 (34, 35). As such, high levels of TWEAK might expand the inflammatory response observed in COVID-19 patients. TSLP is a promoter of allergic inflammation and Th2 responses (36). Indeed, high TSLP levels coincide with a Th2 cytokine profile in our COVID-19 cohort. Our results also indicate a possible role for MMP-1 and MMP-3 in lung injury associated with COVID-19, two matrix metalloproteases implicated in tissue damage underlying other lung diseases (37-39).

## Discussion

During the upcoming winter in the Northern hemisphere, the world will face one of the most challenging public health crises in recent history due to the convergence of influenza and COVID-19. Thus, a better understanding of the clinical and immunopathological characteristics that differentiate both diseases is urgent to guide specific therapeutic approaches for patients with influenza and COVID-19. This includes the selection of adequate antiviral drugs as well as appropriate immunological therapeutics for each case. Unfortunately, whereas our knowledge of the immunopathogenesis of pandemic influenza A(H1N1) has improved over the last decade, resulting in more effective vaccination and pharmacological interventions, the current lack of understanding of the pathobiology of COVID-19 is a barrier to the identification of therapeutic targets for drug and vaccine development. The inevitable co-circulation of influenza viruses and SARS-CoV-2 and the potential scenarios of co-infection with both viral subtypes may further represent an aggravation of the COVID-19 morbidity and mortality. However, we do not know if an over infection with SARS-CoV-2 in patients already infected with influenza viruses would result in worse or better clinical outcomes. The results of the opposite scenario are also speculative. Despite this, it is essential to have reliable indicators that allow differential identification of these conditions, especially in settings of limited resources to perform RT-PCR tests.

Some recent literature reviews have tried to highlight essential differences in the epidemiological characteristics, mechanisms of transmission, and clinical symptoms of patients infected with SARS-CoV-2 and influenza viruses (14, 15). However, these retrospective comparisons carry the risk of biased conclusions due to differences in the genetic background, sociocultural characteristics, and access to medical attention of populations from different regions. Thus, parallel comparisons of influenza and COVID-19 cases in geographical settings with similar health care resources would provide a better perspective of the main differences between these infectious entities. In this context, Mexico is an ideal place to conduct comparative studies between influenza and COVID-19, as this country was the site of origin of the influenza A(H1N1) pdm09 virus (2-4). Since its emergence in 2009, hospitals around Mexico have acquired ample experience in the management of severe cases of this viral infection, which has resulted in progressive decreases in mortality rates over the last ten years (40). On February 28^th^, 2020, Mexico confirmed its first two cases of SARS-CoV-2. Ever since, the epidemiological curve of COVID-19 shows a continuous increase in the number of positive cases, with more than 500,000 cases and 52,000 deaths reported on August 6^th^ of 2020 (41).

Here, we compared the clinical, histopathological, and immune characteristics of patients with pandemic influenza A(H1N1) and COVID-19. One of the most striking findings of our study was that most of the clinical and laboratory parameters routinely evaluated in emergency departments were similar between critically ill influenza and COVID-19 patients, even when some of them can be used to separate moderate from severe COVID-19 and influenza patients. Interestingly, our data reveal that respiratory symptoms are more common during influenza, whereas dry cough and higher frequency of gastrointestinal symptoms are distinctive characteristics associated with SARS-CoV-2 infection. These differences in clinical symptoms may traduce distinct infective capacities of both viruses to affect several organs besides the lungs. In this sense, influenza viruses are thought to be primary respiratory pathogens that rarely cause extrapulmonary manifestations associated with injury directly driven by the virus (42). Meanwhile, it is accepted that SARS-CoV-2 has a broad infective capacity to invade several tissues and organs (43). The expression of the angiotensin I converting enzyme 2 (ACE2), the transmembrane serine protease 2 (TMPRSS2), and 4 (TMPRSS4), furin, and cathepsin L in human organs determine the tissue tropism of SARS-CoV-2. These factors are expressed in the lungs; nonetheless, their expression is even higher at several parts of the upper and lower gastrointestinal tract (44). This might explain the clinical differences observed in our study.

We also found that levels of ALP, ALT, LDH, CPK, procalcitonin, as well as SOFA and APACHE II scores were higher in influenza as compared to both groups of patients with moderate and severe COVID-19. Meanwhile, the PaO2/FiO2 upon arrival was similar in the severe COVID-19 and severe influenza patients. These findings are almost entirely coincident with the results of a recent study evaluating the differences in clinical presentations between Chinese patients with ARDS infected with either SARS-CoV-2 or influenza A(H1N1) (13). In that investigation, the researchers also found that ground-glass opacities were more common in radiological studies of COVID-19 patients, whereas consolidation opacities were more frequent in influenza subjects. Ground-glass opacities are typically associated with an interstitial inflammatory process of the lung, whereas consolidations traduce intra-alveolar exudates (45). Here, we found that the histopathological pattern induced after lung infection with SARS-CoV-2 is mainly characterized by an interstitial inflammatory infiltrate, whereas influenza induces changes compatible with alveolar pneumonia. Together, both studies highlight that influenza and COVID-19 display crucial differences in the histological characteristics of the infected lungs that may also translate into distinctive clinical manifestations.

Furthermore, our study demonstrates that the immune response against SARS-CoV-2 is entirely different from the response against influenza. Indeed, our analyses bring forward a set of immunological markers with the potential to differentiate COVID-19 from influenza successfully. Measuring some of these markers might improve the diagnostic approach and subsequent therapeutic decision for patients with ARDS infected with either SARS-CoV-2 or influenza A(H1N1) pdm09 virus. Also, the differences in the immune profiles detected in the serum and plasma of our cohorts of influenza and COVID-19 patients emphasize the idea that both pathogens elicit distinct host protective responses with non-overlapping effector functions. Although the immune response against SARS-CoV-2 infection is not well comprehended so far, the prevailing paradigm to explain the morbidity and mortality of COVID-19 patients is that SARS-CoV-2 elicits an exuberant immune reaction characterized by a dysregulated production of soluble immune mediators. This phenomenon, known as “cytokine storm,” is thought to be responsible for mediating tissue injury in patients with COVID-19 that progress to severe illness (22, 46-48).

The immune receptors that recognize the viral infection and initiate the immune responses against SARS-CoV-2 are unknown. As this virus is genetically related to SARS-CoV-1, it is presumed that both viruses share mechanisms of infection. In this sense, SARS-CoV-1 is recognized by the toll-like receptors (TLR) TLR3 and TLR4, which induce an immune reaction via MyD88 and TRIF pathways (49, 50). Furthermore, SARS-CoV-1 triggers the production of IL-1β through the activation of the inflammasome (51). It is also possible that SARS-CoV-2 activates the inflammasome, as high levels of IL-1β have been observed in COVID-19 patients (52). Other immune mediators exaggeratedly produced in response to SARS-CoV-2 include IL2, IL-6, IL7, IL10, G-SCF, CXCL10, CCL2, CCL3, and TNF-α (9, 22, 23). Similar immune signatures were detected in our cohort of COVID-19 patients.

Meanwhile, the pathogenicity and virulence of the influenza A(H1N1) pdm09 virus are due to acquired properties contributing to alter the regulation of inflammatory responses and evade antiviral immunity. In previous studies, we have described that pandemic, but not seasonal influenza A strains, can downregulate the expression of the suppressors of cytokine signaling 1 (SOCS-1) and increase the production of IL-6, IL-8, TNF-α, IL-10, CCL3, CCL4, and CCL5 in experimental infection assays of human lung A549 epithelial cells and human macrophages (53). Levels of IL-6, IL-8, TNF-α, and CCL3 were also increased in our cohort of influenza patients, validating our previous observations. The influenza A(H1N1) pdm09 also suppresses the expression of the retinoid-inducible gene I (RIG-I) and induces lower levels of type I interferons in human macrophages and human lung epithelial cells, as compared to seasonal influenza A strains. In this sense, it is possible that blocking type I interferon responses might be a strategy of SARS-CoV-2 to evade antiviral immune mechanisms, as we found very low induction of plasma IFN-α and IFN-β in both influenza and COVID-19 patients. These findings are in line with previous studies showing alterations of type I interferons in critically ill COVID-19 patients (54).

Notably, despite the dysregulated production of other immune mediators, an ample range of immune cell subtypes are depleted from the circulation of patients with severe SARS-CoV-2 infection. These cells include monocytes, dendritic cells, CD4+ T cells, CD8+ T cells, B cells, and NK cells (55). Furthermore, the few adaptive lymphocytes that remain in the blood express markers of functional exhaustion (24). These data suggest that severe COVID-19 is a state of immunosuppression similar to the known sepsis-induced immunosuppression (56). Notably, a recent study by Remy and collaborators has shown that the immunosuppression observed in COVID-19 is even more profound than in critically ill patients with sepsis of other causes (21). In such investigation, functional assays demonstrated that the production of IFN-γ by T cells isolated from the peripheral blood of COVID-19 patients and stimulated with anti-CD3/anti-CD28 antibodies was impaired as compared with T cells from healthy individuals and septic patients. Furthermore, a reduced production of TNF-α by stimulated monocytes from COVID-19 patients was noticed. These findings led the researchers to propose that the primary immune mechanism underlying the morbidity and mortality of COVID-19 is immunosuppression rather than hyperinflammation.

In this context, our study may provide additional evidence useful to clarify these controversies. Based on our results and previous investigations, we propose that hyperinflammation and immunosuppression are not mutually exclusive in COVID-19. First, our data showed some indirect readouts of immunosuppression in individuals infected with SARS-CoV-2. For instance, we found that TNF-α levels were lower in the serum of COVID-19 patients as compared to influenza patients. This coincides with the limited capacity of monocytes from COVID-19 patients to produce TNF-α upon stimulation described by Remy et al. (21). We also observed lower plasma levels of the macrophage activation marker sCD163, although macrophages infiltrating the lungs of COVID-19 patients expressed several cytokines. Furthermore, we found low levels of IL-2 and APRIL/TNFSF13 (two immune mediators crucial for T-cell and plasma cell survival), as well as sCD30 (a marker of lymphocyte activation) in the circulation of COVID-19 but not influenza patients. Similarly, we observed a lack of lymphocytes in the inflammatory infiltrates found in lung autopsy specimens from patients that died of COVID-19. These findings may reflect a depletion of activated lymphocytes and monocytes from the circulation during SARS-CoV-2 infection and poor recruitment of lymphocytes to the lungs.

At the same time, we have described that an exacerbated polyfunctional immune response prevails in the circulation of COVID-19 patients. Such a response is characterized by higher levels of Th1 as well as Th2 cytokines as compared to influenza patients. Conversely, although influenza subjects also display elevated levels of some inflammatory mediators, these individuals may have enough regulatory mechanisms that counteract the detrimental effects of hyperinflammation. The higher levels of IL-1RA observed here in influenza patients as compared to COVID-19 subjects well exemplify this. Furthermore, we found higher serum levels of the C-X-C motif chemokine ligand 17 (CXCL17), a mucosal chemokine with anti-inflammatory properties, in influenza but not COVID-19 patients (unpublished findings). In addition, the serum cytokine pattern of COVID-19 resembles the inflammatory profile of rheumatoid arthritis patients with interstitial lung disease (57), and the polyfunctional inflammatory response of the cytokine release syndrome (CRS) that occurs after chimeric antigen receptor (CAR) T-cell therapy (58). Immunosuppression and hyperinflammation are also a hallmark of both of these conditions.

Notably, the higher levels of Th2 cytokines, particularly IL-4 and IL-5, might inhibit Th1 protective antiviral responses in COVID-19 patients. Thus, our data indicate that a lack of immune balance of the type of effector response is another crucial determinant of the collapse of the host protective immunity against SARS-CoV-2. This Th2 biased response may generate interstitial infiltrates of Th2 cells, neutrophils, eosinophils, and type 2 innate lymphoid cells, mediating lung inflammation, and tissue damage. In fact, critically ill COVID-19 patients that have undergone lung histopathological analyses or subsequent CT scans showed interstitial lung infiltrates, some of which resemble several forms of progressive interstitial lung disease like cryptogenic organizing pneumonia and non-specific interstitial pneumonia (9, 59-61). Here, we also observed interstitial inflammation and expression of IL-4 in the lungs of COVID-19 patients but not influenza subjects. These deleterious effects of Th2 responses in the lungs could also explain the abnormalities in lung function, and potential progression to pulmonary fibrosis observed in more than 45 % of COVID-19 patients discharged from hospitals (62), particularly in older patients, which exhibit a higher rate of post-COVID-19 lung fibrosis. Hence, it would be of great interest to characterize the cytokine profile of COVID-19 patients that subsequently develop any form of interstitial lung disease, as they would benefit from specific and anti-fibrotic therapeutics.

We propose that ideal immune therapeutics for COVID-19 should be directed not only to blocking or enhancing specific immune signaling pathways to counteract hyperinflammation or reverting immunosuppression, but rather to re-establish a convenient immune balance that promotes protective immunity. Under the light of this hypothesis, several immune mediators and immune cell subsets could be targeted for therapeutic purposes. For instance, type 2 innate lymphoid cells (ILC2s) have been identified as the leading producers of Th2 cytokines in the lungs, contributing to potent allergen-induced airway inflammation even in lymphopenic hosts (63). Thus, ILC2s may constitute novel targets to inhibit Th2 responses in COVID-19 patients. The potential pathogenic effects of Th2-biased responses in COVID-19 may also be counteracted with monoclonal antibodies. For instance, dupilumab, a monoclonal antibody against IL-4, has been safely used in patients with atopic dermatitis and COVID-19, without increased risk of severe complications of the infection, and even some patients receiving dupilumab that later acquired the infection with SARS-CoV-2 did not show respiratory symptoms (64-66). Finally, TSLP could be another target to inhibit Th2 responses in COVID-19 patients, as this molecule promotes allergic inflammation (36), and indeed, high levels of TSLP were observed in our cohort of COVID-19 but not influenza subjects.

### Limitations

A limitation of our study is that we did not recruit patients infected with seasonal influenza virus subtypes. Thus, our observations are only useful to distinguish between influenza A(H1N1) pdm09 and SARS-CoV-2 infection. The clinical and immunological characteristics of SARS-CoV-2 and seasonal influenza have been compared in a recent study by Mudd et al., which is under review (67). In such a study, researchers found that COVID-19, as compared to seasonal influenza, is characterized by lower mean cytokine levels in serum. Conversely, we found that cytokine levels were higher in COVID-19 patients than in individuals with pandemic influenza A(H1N1). These discrepancies are probably related to variations in the virulence and capacity to induce inflammatory immune responses of seasonal and pandemic influenza virus subtypes. Finally, another limitation of our study is that we did not measure cytokine levels in serial serum/plasma samples from our two cohorts of influenza and COVID-19 patients. Thus, future investigations should compare differences in the kinetics of immune responses against both diseases. Despite this, our study provides important insights into the differences between the two most important respiratory pathogens that have caused pandemics of international concern in recent years.

## Conclusions

In conclusion, our results demonstrate significant differences in the immune responses elicited after SARS-CoV-2 and influenza A(H1N1) pdm09 virus. Our data support the use of specific clinical characteristics, laboratory parameters, and immunological markers to differentiate SARS-CoV-2 infection from pandemic influenza A(H1N1). These data may also contribute to the discovery of novel therapeutic targets to counteract harmful immune mechanisms underlying the immunopathology of COVID-19 and influenza.

## Methods

### Study design and participants

We conducted a prospective cohort study in patients with an acute respiratory illness that attended the emergency department of the Instituto Nacional de Ciencias Médicas y Nutrición Salvador Zubirán (INCMNSZ), and the Instituto Nacional de Enfermedades Respiratorias Ismael Cosío Villegas (INER) in Mexico City. Individuals requiring hospital admission that tested positive for SARS-CoV-2 infection by real-time polymerase chain reaction (RT-PCR) in swab samples, bronchial aspirates (BA), or bronchoalveolar lavage (BAL) specimens were eligible. Detection of SARS-CoV-2 was performed by RT-PCR, as previously described (68). Briefly, viral RNA was extracted from clinical samples with the MagNA Pure 96 system (Roche, Penzberg, Germany). The RT-PCR reactions were performed in a total volume of 25μL, containing 5μL of RNA, 12.5μL of 2 × reaction buffer provided with the Superscript III one-step RT-PCR system with Platinum Taq Polymerase (Invitrogen, Darmstadt, Germany; containing 0.4 mM of each deoxyribose triphosphates (dNTP) and 3.2 mM magnesium sulfate), 1μL of reverse transcriptase/ Taq mixture from the kit, 0.4μL of a 50 mM magnesium sulfate solution (Invitrogen), and 1μg of nonacetylated bovine serum albumin (Roche). Primer and probe sequences, as well as optimized concentrations, are shown in Supplemental Table 1. All oligonucleotides were synthesized and provided by Tib-Molbiol (Berlin, Germany). Thermal cycling was performed at 55 °C for 10 min for reverse transcription, followed by 95 °C for 3 min and then 45 cycles of 95°C for 15 s, 58°C for 30 s.

Individuals positive for SARS-CoV-2 infection by RT-PCR were further categorized into two groups: a) moderate COVID-19 group (n=10), that included patients with respiratory symptoms that did not require MV; and b) severe COVID-19 group (n=24), consisting of patients requiring invasive MV and admission to the ICU. Our comparative cohort included patients with influenza-like illness (ILI) that attended to the INER in Mexico City during the immediately preceding 2019/2020 flu season. Individuals with confirmed influenza A(H1N1) pdm09 virus infection that progressed to acute respiratory distress syndrome (ARDS), requiring MV and admission to the ICU were included. ILI was defined as an acute respiratory illness with a measured temperature of ≥ 38 °C and cough, with onset within the past ten days. These subjects were first screened for influenza A virus infection using the Fuji dri-chem immuno AG cartridge FluAB kit (Fujifilm Corp, Tokyo, Japan) rapid influenza diagnostic test (RIDT) in fresh respiratory swab specimens. In positive cases, further molecular characterization of the causative influenza A virus subtype was assessed by RT-PCR. None of the patients enrolled in the study had human immunodeficiency virus (HIV) infection.

### Data retrieval

Microsoft Excel (MS Excel 365) was used for data collection of the epidemiological and clinical information. Clinical and demographic data were retrieved from the medical records of all participants. These data included age, gender, anthropometrics, comorbidities, symptoms, triage vital signs, the severity of illness scores at admission (Sequential Organ Failure Assessment (SOFA), and Acute Physiology And Chronic Health Evaluation II (APACHE II)), as well as initial laboratory tests. Initial laboratory tests were defined as the first test results available (typically within 24 hours of admission) and included white blood cell counts, liver and kidney function, geometric parameters at admission, and other biomarkers.

### Cytokine determinations

Peripheral blood samples were obtained from all participants at hospital admission. Serum levels of different cytokines, chemokines, growth factors, and other immune mediators were determined by Luminex assays using the Luminex platform Bio-Plex Multiplex 200 (Bio-Rad Laboratories, Inc., Hercules, CA, USA). Serum samples from 13 healthy donors were used as controls. The immune mediators that were quantified are listed as follows: IFN-α, interferon-alpha, IFN-β; interferon-beta; IFN-γ, interferon-gamma; TNF-α, tumor necrosis factor-alpha; IL-1β, interleukin 1beta; IL-1RA, interleukin 1 receptor antagonist; IL-2, interleukin 2; IL-4, interleukin 4; IL-5, interleukin 5; IL-6, interleukin 6; IL-7, interleukin 7; IL-8, interleukin 8; IL-9, interleukin 9; IL-10, interleukin 10; IL-12 (p40), interleukin 12 p40 subunit; IL-12p70, interleukin 12 p70 subunit; IL-13, interleukin 13; IL-15, interleukin 15; IL-17A, interleukin 17A; IL-26, interleukin 26; IL-32, interleukin 32; CXCL10, C-X-C motif chemokine ligand 10, CCL2, C-C motif chemokine ligand 2; CCL3, C-C motif chemokine ligand 3; CCL4, C-C motif chemokine ligand 4; CCL5, C-C motif chemokine ligand 5; CCL11, C-C motif chemokine ligand 11; G-CSF, granulocyte colony-stimulating factor; bFGF, basic fibroblast growth factor; PDGF-BB, platelet-derived growth factor bb; VEGF, vascular endothelial growth factor; APRIL/TNFSF13, A proliferation-inducing ligand/tumor necrosis factor ligand superfamily member 13; BAFF/TNFSF13B, B-cell activating factor/tumor necrosis factor ligand superfamily member 13B; sCD30/TNFRSF8, soluble CD30/ tumor necrosis factor ligand superfamily member 8; sCD163, soluble CD163; chitinase 3/like1; gp130/sIL-6Rβ, glycoprotein of 130 kDa/soluble IL-6 receptor beta; sIL-6Rα, soluble IL-6 receptor alpha; MMP-1, metalloprotease 1; MMP-2, metalloprotease 2; MMP-3, metalloprotease 3; osteocalcin; ostepontin; pentraxin-3; sTNF-R1, soluble tumor necrosis factor receptor 1; sTNF-R2, soluble tumor necrosis factor receptor 2; TSLP, thymic stromal lymphopoietin; TWEAK/TNFSF12, tumor necrosis factor-like weak inducer of apoptosis/tumor necrosis factor ligand superfamily member 12.

### Histological analysis and immunohistochemistry

Formalin-fixed and paraffin-embedded lung autopsy specimens from patients who died of influenza or COVID-19 (N=2 patients per group) were obtained from the Pathology Department of the INER. Sections of 3-5μm were processed for hematoxylin-eosin (H&E) staining for histopathological analysis. For immunohistochemistry (IHQ), lung sections were mounted on silane-covered slides, deparaffinized in xylenes, and hydrated with a series of graded alcohol-to-water dilutions. The endogenous peroxidase was blocked with 3% hydrogen peroxide for 30 minutes. Sections were incubated overnight at room temperature with optimal dilutions (1:100) of the following antibodies: anti-IFN-γ (Anti-Interferon gamma antibody, ab9657, Abcam, UK), anti-IL-1β (IL-1β Antibody (H-153): sc-7884, Santa Cruz Biotechnology Inc., Santa Cruz, CA), anti-IL-4 (IL-4 Antibody (OX81): sc-53084, Santa Cruz Biotechnology Inc., Santa Cruz, CA), and anti-IL-17A (Anti-IL-17 antibody (ab91649), Abcam, UK). Secondary biotinylated antibodies labeled with peroxidase were added, and those attached were revealed with diaminobenzidine (DAB) for 5 minutes (MACH 1 Universal HRP-Polymer Detection Kit, Biocare Medical, LLC). Slides were counter-stained with hematoxylin.

### Statistical analysis

Descriptive statistics were used to characterize the study population clinically. Frequencies and proportions were calculated for categorical data. Means, medians, standard deviations (SD), interquartile ranges (IQR), and 95% confidence intervals were used for continuous variables. Differences in categorical variables between groups were assessed by the Fisher exact or Chi-square test. For comparisons of continuous variables between two groups, we used the Mann-Whitney U test. For differences of continuous between more than two groups, we used the Kruskal-Wallis test with post hoc Dunn’s test. Multiple linear regression analyses using Spearman rank correlation coefficients were used to determine correlations between continuous variables. ROC curves were constructed to estimate the diagnostic utility of different variables to differentiate between participant groups in terms of their area under the curve (AUC). The prognostic value of the different clinical and immunological parameters expressed in terms of odds ratio (OR) values for adverse outcomes (intubation, death) was estimated using binomial logistic regression analyses.

Principal component analyses (PCAs) were conducted to analyze how the study participants clustered together according to the interplay between their clinical and immunological characteristics. Furthermore, a Linear Discriminant Analysis (LDA) without and with “leave-one-out” type cross-validation was performed to assess whether the linear combination of different variables allowed differentiating individuals according to their diagnosis. The variables included were AST, ALT, LDH, ALP, procalcitonin, SOFA, IL-β, IL-1RA, IL-2, IL-4, IL-5, IL-7, IL-12, IL-13, IL-17A, TNF-α, CCL3, CCL11, G-CSF, and VEGF. A Wilks ’Lambda test was performed to evaluate the discriminatory power of each variable in the LDA. Variables were transformed to log10 to meet the LDA assumptions and were scaled to prevent the scale of each variable from influencing the analysis results. Individuals with missing data were omitted from PCA and LDA analyses. All analyses were conducted using GraphPad Prism 8 (La Jolla, CA), R Statistical Software (Foundation for Statistical Computing, Vienna, Austria) packages Factoextra and MASS, and Python packages pandas v0.23.4 and seaborn v0.10.1. Specific analysis tests are also mentioned in figure legends. P values ≤0.05 were considered as significant: *p≤0.05, **p≤0.01, ***p≤0.001, ****p≤0.0001.

### Study approval

The Institutional Review Boards of the Instituto Nacional de Ciencias Médicas y Nutrición Salvador Zubirán (INCMNSZ, approval number: 3349) and the Instituto Nacional de Enfermedades Respiratorias Ismael Cosío Villegas (INER, approval number: B28-16 and B09-20) in Mexico City approved the study.

All participants or their legal guardians provided written informed consent in accordance with the Declaration of Helsinki for Human Research. Clinical samples were managed according to the Mexican Constitution law NOM-012-SSA3-2012, which establishes the criteria for the execution of clinical investigations in humans.

## Data Availability

The databases used in the current study are available from the corresponding author on reasonable request

## Author contributions

Designed of the research study: JC-P, TR-R, SAK, AZ, and JZ. Recruited patients: JC-P, TR-R, MS-V, DH-G, EM-G, ES, JM-R, JB-R, HV-R, G-CS, N-AP, GH, JG, LM-H, LP-B, GD-C, and CH-C. Retrieved clinical data: JC-P, TR-R, MS-V, DH-G, N-AP, GH, LM-H, CH-C, AH-M, LO. Collected and processed blood samples: JC-P, LJ-A, AC-L, TR-R, GR-M, EM-G, NA-P, GH, CM-M, AD, and LM-H. Obtained and processed lung autopsy specimens: CS-L, CS-G, and CL. Conducted cytokine determinations: L-JA, AC-L, GR-M, and EM-G. Conducted histological and immunohistochemistry analyses: JC-P, CS-L, and CS-G. Performed statistical analyses of the data: JC-P, EC-P, YB-M, and MM-S. Provided reagents: LJ-A, TR-R, CS-L, JG, LM-H, LP-B, GD-C, CC-G, JS-H, PS-D, JR, EG-L, CH-C, SAK, AZ, and JZ. Discussed the manuscript: JC-P, TR-R, SAK, AZ, and JZ. Drafted the manuscript: JC-P, SAK, AZ, and JZ. All authors read and approved the final version of the manuscript.

## Acknowledgments

JAPC was supported by the National Council of Science and Technology of Mexico to achieve (CONACyT) his PhD degree (CONACyT-CVU 737347). The current study was supported by institutional research funds of INER and by research contracts: SECTEI/050/2020, Secretaría de Ciencia, Tecnología e Innovación de la Ciudad de México (SECTEI CDMX); FORDECyT/10SE/2020/05/14-06 and FORDECyT/10SE/2020/05/14-07 from the Fondo Institucional de Fomento Regional para el Desarrollo Científico y Tecnológico y de Innovación (FORDECyT) of the CONACyT.

